# Care-Related Determinants of Adverse Outcomes among Low-Birth-Weight Neonates: Evidence from Newborn Unit Practice and Provider Perspectives in Kenya

**DOI:** 10.64898/2026.07.13.26357939

**Authors:** Judy Cheptoo, Vincent K. Mukthar, Morris S. Shisanya

**Affiliations:** Department of Nursing, Kabarak University, Private Bag 20157, Nakuru, Kenya; Department of Nursing, Egerton University, P.O. Box 536-20115, Egerton, Kenya; Department of Community Health Nursing, School of Nursing, Kibabii University, P.O. Box 1699-50200, Bungoma, Kenya

**Keywords:** low-birth-weight neonates, newborn unit, nursing care processes, warm chain, infection prevention, referral coordination, drug and equipment shortage, quality improvement, provider perspectives, mixed methods

## Abstract

**Context:** Survival of low-birth-weight (LBW) neonates depends heavily on modifiable nursing care processes at the bedside, making the newborn unit a decisive site for improvement. Evidence linking measurable care-process conditions to outcomes, and the provider experience that explains them, remains limited in Kenyan county referral settings.

**Aim:** To examine the care-related determinants of severe adverse outcomes among LBW neonates and the provider perspectives that explain them, framed for newborn-unit nursing practice and quality improvement.

**Methods:** A convergent mixed-methods design was applied at Kericho County Referral Hospital. Quantitatively, 169 LBW neonate-mother pairs were analysed; care-process indicators (skilled personnel at admission, warm-chain maintenance, shortage of essential drugs/feeds, and referral/outborn status) were related to a composite severe adverse outcome using Pearson chi-square tests and crude odds ratios (OR) with 95% confidence intervals (CI). Qualitatively, key-informant interviews with newborn-unit providers were analysed thematically and coded to care-process themes; strands were integrated for practice.

**Findings:** A severe adverse outcome occurred in 136/169 neonates (80.5%). Skilled personnel (94.7%) and warm-chain practices (92.9%) were near-universal, whereas 58.6% of neonates faced shortages of essential drugs/feeds and 49.1% were referred (outborn). Shortage of essential drugs/feeds (OR = 2.26, 95% CI [1.04, 4.90], p = .036) and referral/outborn status (OR = 2.25, 95% CI [1.01, 5.00], p = .043) were significantly associated with higher odds of a severe outcome. Warm-chain care was statistically associated but in a counterintuitive direction (OR = 4.81, p = .006), consistent with confounding by indication, while skilled-personnel availability was not associated (p = .834). Provider narratives converged on seven care-process themes: staffing and workload, warm-chain maintenance, infection prevention, drug and equipment availability, referral coordination, monitoring and documentation, and caregiver/transport barriers around the first hour of care.

**Conclusion:** Care-related conditions—commodity supply, referral readiness, thermal care, infection prevention, and monitoring—are clinically modifiable levers that shape whether vulnerable LBW neonates stabilize or deteriorate, even where they do not all retain independent statistical significance after adjustment.

**Recommendations:** Newborn units should protect nurse staffing norms, secure consistent supply of essential neonatal commodities, standardize pre-referral stabilization and thermal-care protocols, and strengthen structured monitoring and documentation, supported by competency-based training and county-level policy.

## Introduction

Low birth weight (below 2,500 g) remains one of the strongest correlates of neonatal morbidity and mortality worldwide, and its burden is concentrated in low- and middle-income countries. Globally, an estimated one in seven newborns is born with low birth weight, and these infants account for a disproportionate share of neonatal deaths from complications such as respiratory distress, sepsis, hypothermia, and hypoglycaemia (Blencowe et al., 2019; WHO, 2024). In sub-Saharan Africa, and Kenya specifically, the neonatal mortality rate has declined more slowly than post-neonatal mortality, leaving the newborn period as the critical bottleneck to further reductions in under-five mortality (Kenya National Bureau of Statistics & ICF, 2023).

Whereas the biological vulnerability of LBW neonates is largely fixed at birth, their trajectory after admission is shaped substantially by the quality and readiness of the care they receive. Bedside nursing processes—thermal protection through the warm chain, infection prevention, feeding support, structured monitoring, timely recognition of deterioration, and prompt escalation—together determine whether a small or preterm neonate stabilizes or progresses to a severe adverse outcome (WHO, 2022). These processes are, in principle, modifiable: they respond to staffing levels, commodity supply, protocol adherence, and system readiness rather than to fixed patient characteristics. This makes the newborn unit a decisive site for nursing-led quality improvement.

Yet evidence that links measurable, care-related conditions to neonatal outcomes—and that pairs those associations with the lived experience of the providers who deliver care—remains limited in Kenyan county referral settings. Care-process factors are frequently discussed narratively but seldom quantified alongside outcomes, and provider perspectives that could explain the mechanisms behind the numbers are rarely integrated. Understanding both the statistical signal and the practice reality is essential if newborn-unit nurses, unit managers, educators, and quality-improvement teams are to target the conditions most amenable to change.

The complications that drive severe outcomes in this population—respiratory distress, neonatal sepsis, hypothermia, and hypoglycaemia—are strongly time- and care-sensitive. Preterm and low-birth-weight neonates lose heat rapidly, tolerate feeds poorly, and are highly susceptible to hospital-acquired infection, so that even short lapses in thermal protection, hygiene, feeding support, or monitoring can precipitate deterioration (Chawanpaiboon et al., 2019; Seale et al., 2014). In high-mortality settings, a substantial share of neonatal deaths is attributable not to the absence of known interventions but to gaps in their reliable delivery at the bedside—what has been termed the quality gap in newborn care (Moxon et al., 2015). For LBW neonates, therefore, the newborn unit is not merely a place of observation but an active determinant of survival.

Nursing staff are the constant presence in that environment. They maintain the warm chain, enforce infection-prevention practice, support feeding and kangaroo mother care, monitor for early signs of deterioration, and coordinate escalation and referral. Evidence from Kenyan hospitals shows that the availability, knowledge, and workload of newborn-unit nurses are tightly linked to whether essential newborn-care practices are actually performed (English et al., 2017; Gathara et al., 2020). Because these care-process conditions are modifiable, they represent the most tractable targets for improving outcomes among neonates whose biological risk is already established at birth. Quantifying which conditions matter—and understanding, from providers themselves, why—is the necessary first step toward focused, nursing-led quality improvement.

This paper therefore focuses on the care-related determinants of severe adverse outcomes among LBW neonates admitted to a county referral hospital in Kenya, integrating quantitative associations between care-process indicators and outcomes with provider narratives that explain how practice conditions translate into neonatal risk.

## Statement of Significance

Most published determinant analyses of LBW neonatal outcomes emphasise maternal and neonatal biological factors, which are difficult to modify at the point of care. By contrast, this study isolates care-process conditions—commodity supply, referral readiness, thermal care, infection prevention, staffing, and monitoring—that fall squarely within the remit of newborn-unit nursing and facility management. Pairing measurable care-process associations with provider accounts gives nurses and quality-improvement teams both the evidence and the mechanism needed to prioritise bedside and system changes. The findings translate directly into practice, education, and policy levers for improving survival of vulnerable neonates in resource-constrained referral units.

## Aim of the Study

To examine the care-related determinants of severe adverse outcomes among low-birth-weight neonates and the provider perspectives that explain them, to identify modifiable nursing-practice and quality-improvement targets. The specific objectives were: (a) to describe the frequency of key care-process conditions in the newborn unit; (b) to assess the association between these care-process conditions and severe adverse outcomes; and (c) to explore, through provider key-informant interviews, how care-process conditions shape neonatal outcomes at the bedside and system level.

## Operational Definitions

### Low-birth-weight (LBW) neonate

A live-born neonate with a birth weight below 2,500 g, admitted to the newborn unit.

### Severe adverse outcome

A composite indicator, coded present when a neonate experienced any of respiratory distress, neonatal sepsis, hypothermia, hypoglycaemia, prolonged newborn-unit stay (≥7 days), or death during admission.

### Warm-chain maintenance

Documented use of thermal-protection practices (e.g., immediate drying, skin-to-skin/kangaroo mother care, and incubator or warmer use) to prevent heat loss.

### Skilled personnel at admission

Presence of a health worker trained in newborn care at the time the neonate was received in the unit.

### Shortage of essential drugs/feeds

A documented gap in the availability of essential neonatal medicines or feeds during the admission.

### Referral / outborn status

The neonate was delivered at, or transferred from, another facility rather than born within the study hospital.

### Care-process indicators

Measurable conditions of care delivery (staffing, thermal care, infection prevention, commodity supply, referral coordination, and monitoring/documentation) distinguished from fixed maternal and neonatal biological factors.

## Subjects and Methods

### Study Design and Setting

A convergent mixed-methods design was used, combining a hospital-based, cross-sectional quantitative strand with a qualitative key-informant strand. The study was conducted in the newborn unit of Kericho County Referral Hospital (KCRH), a public referral facility in the South Rift region of Kenya that receives both inborn deliveries and outborn referrals from lower-level facilities across the county.

### Population and Sampling

The quantitative sample comprised 169 low-birth-weight neonate-mother pairs admitted to the newborn unit during the study period, enrolled consecutively. For the qualitative strand, nine healthcare providers involved in newborn care were purposively selected to capture a range of cadres and experience: one paediatrician, five nurses, two clinical officers, and one unit manager, with between two and ten years of neonatal experience. Purposive selection prioritised providers with direct, sustained involvement in the care of LBW neonates.

### Care-Related Variables

The care-process indicators examined were skilled-personnel availability at admission, warm-chain maintenance, shortage of essential neonatal drugs or feeds, and referral/outborn status. These were selected because they are directly observable at the point of care, are shaped by nursing practice and facility readiness rather than fixed patient biology, and correspond to the domains raised by providers in the qualitative strand (staffing and workload, thermal care, infection prevention, commodity supply, referral coordination, and monitoring and documentation).

### Outcome Measure

The primary outcome was a composite severe adverse outcome (defined above), consistent with the parent study, reflecting the clinically important events that newborn-unit care seeks to prevent.

### Data Collection

Quantitative data were abstracted from admission and clinical records using a structured tool. Qualitative data were collected through semi-structured key-informant interviews (30–45 minutes each) guided by an interview schedule covering perceived determinants, equipment and staffing adequacy, timeliness of care, referral effectiveness, barriers and delays, and recommendations. Interviews were conducted in private, after informed consent, and were recorded and transcribed.

### Data Analysis

Quantitative analysis was descriptive-analytic rather than a full adjusted model, in keeping with the nursing-practice focus of this paper and to avoid duplicating the parent determinants analysis. Frequencies were computed for each care-process indicator. Associations between care-process conditions and the severe adverse outcome were tested using Pearson chi-square tests (Fisher’s exact test where expected cell counts were small), with crude odds ratios and 95% confidence intervals. Statistical significance was set at p < .05. Qualitative transcripts were analysed thematically following a six-phase approach: familiarisation, coding, theme development, review, definition, and reporting. Codes were organised into care-process themes; an audit trail and reflexivity notes were maintained. The two strands were then integrated to show how measurable care conditions and provider mechanisms align.

### Ethical Considerations

Ethical approval was granted by the Kabarak University Research Ethics Committee (KUREC), approval number KUREC-170226 (Ref. KABU01/KUREC/001/17/02/26), valid 19 February 2026 to 19 February 2027. A research licence was obtained from the National Commission for Science, Technology and Innovation (NACOSTI), licence number NACOSTI/P/26/4186446 (Ref. 625542), issued 3 April 2026. Institutional authorisation was obtained from the hospital administration. Informed consent was obtained from mothers/guardians and from key informants; participation was voluntary and confidential, and neonatal records were de-identified.

## Results

### Provider profile

Nine key informants participated (Table 1), spanning paediatric, nursing, clinical-officer, and unit-management cadres, with two to ten years of newborn-unit experience. Nurses formed the largest group, consistent with their central role in continuous bedside care of LBW neonates.

**Table 1:** Key-Informant (Provider) Profile (n = 9)

| **Cadre** | **Role in newborn care** | **n** | **Experience (years)** |
| --- | --- | --- | --- |
| Paediatrician | Clinical leadership; management of complications | 1 | ~10 |
| Nurse | Continuous bedside care; thermal care, IPC, feeding, monitoring, KMC support | 5 | 2–10 |
| Clinical officer | Admission assessment; prescribing; stabilization | 2 | 3–7 |
| Unit manager | Staffing, supplies, and unit coordination | 1 | ~8 |
| **Total** |  | **9** | **2–10** |

*Note.* Cadres and experience derived from purposive key-informant sampling; nurses provided the majority of continuous bedside care. IPC = infection prevention and control; KMC = kangaroo mother care.

### Care-process profile

Among the 169 neonates, a severe adverse outcome occurred in 136 (80.5%). Skilled personnel at admission (94.7%) and warm-chain practices (92.9%) were near-universal, whereas shortages of essential neonatal drugs or feeds affected 58.6% of neonates, and 49.1% were referred (outborn) from another facility (Table 2).

**Table 2:** Care-Process Indicators (N = 169)

| Care indicator | n (%) |
| --- | --- |
| Skilled personnel available at admission | 160 (94.7) |
| Warm-chain practices maintained | 157 (92.9) |
| Shortage of essential neonatal drugs/feeds | 99 (58.6) |
| Referred from another facility (outborn) | 83 (49.1) |

Note. Values are n (%); N = 169.

### Association with severe adverse outcome

Shortage of essential drugs/feeds (OR = 2.26, 95% CI [1.04, 4.90], p = .036) and referral/outborn status (OR = 2.25, 95% CI [1.01, 5.00], p = .043) were each significantly associated with higher odds of a severe outcome (Table 3). Warm-chain maintenance was also statistically associated (χ^2^ = 7.63, p = .006), but in a counter-intuitive direction: severe outcomes were more common among neonates who received warm-chain care, most plausibly reflecting confounding by indication, because the smallest and sickest neonates are precisely those preferentially given intensive thermal support. Skilled-personnel availability was not associated with the outcome (p = .834). These are unadjusted associations and should be interpreted cautiously, given the small non-severe group (n = 33) and confounding by indication.

**Table 3:**
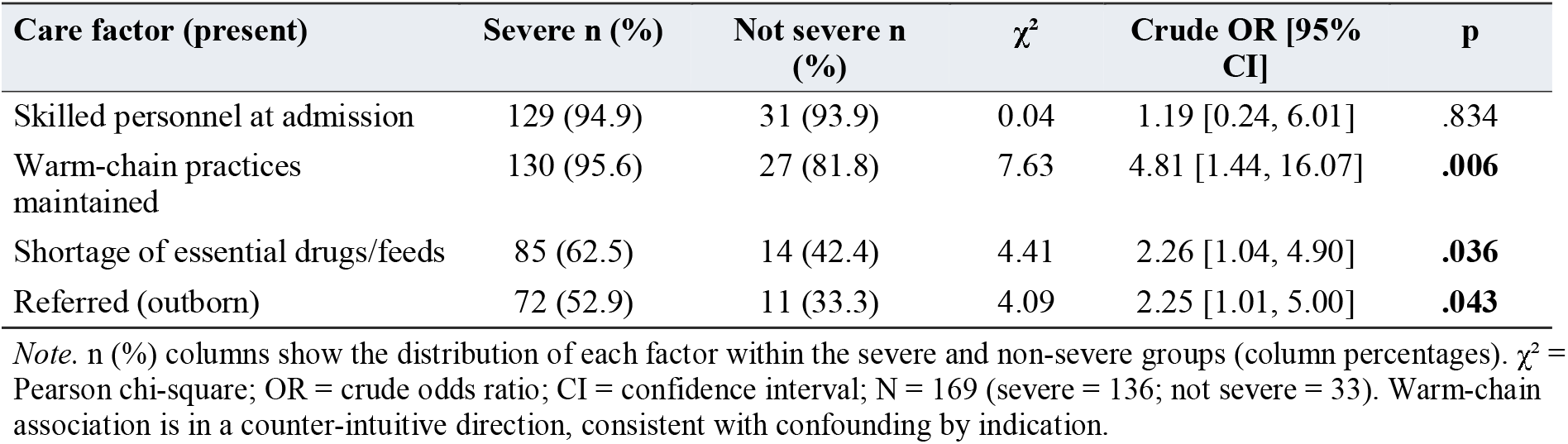
Association of Care-Process Factors With Severe Adverse Outcome.

### Provider Perspectives: Care-Process Themes

Thematic analysis of the key-informant interviews yielded seven care-process themes that explain how practice conditions translate into neonatal risk (Table 4). Interviews were conducted with nine key informants—one pediatrician, five nurses, two clinical officers, and the newborn-unit manager—cited below as KII1 through KII9. Representative verbatim accounts are presented for each theme; minor edits for readability are shown in brackets and omissions as ellipses.

**Table 4:** Care-Process Themes, Subthemes, and Representative Provider Accounts.

| Theme | Subthemes | Representative account (provider) |
| --- | --- | --- |
| Staffing and workload | Understaffing; long hours; reduced productivity | “...you may be monitoring very sick babies, admitting new ones, and assisting mothers with feeding at the same time.” (KII2) |
| Warm-chain maintenance | Delayed skin-to-skin; incubator shortage; heat loss during transfer | “Babies lose heat during transfer from labour ward, theatre, or lower facilities.” (KII6) |
| Infection prevention | Congestion; limited spacing; decontamination gaps | "...it becomes difficult to separate infected and non-infected babies properly." (KII4) |
| Drug & equipment availability | CPAP shortage; inconsistent drug supply; cost | "...when admissions are many, the equipment is not always enough for the number of babies." (KII1) |
| Referral coordination | Unclear referral system; few ambulances; poor pre-referral stabilization | "A small baby should not be transported cold." (KII9) |
| Monitoring & documentation | Slow triage-to-admission; constrained monitoring | "...when one nurse has many babies, it becomes difficult to check everything on time." (KII3) |
| Caregiver & transport barriers | Distance; cultural beliefs; delayed recognition of danger signs | "Some families wait for permission from the father or elder. Transport cost is also a factor." (KII7) |

*Note.* Themes derived from thematic analysis of nine key informant interviews; representative accounts are verbatim (lightly edited for readability).

### Staffing and Workload

> Providers described chronic understaffing that lengthened working hours and strained bedside care.
>
> *“At night, the workload is heavier because fewer staff are on duty. You may be monitoring very sick babies, admitting new ones, and assisting mothers with feeding at the same time*.*”* (KII2, nurse)

### Warm-Chain Maintenance

Thermal care was undermined by heat loss during transfer, incubator shortages, and interruptions to Kangaroo Mother Care.

> *“Warm chain is one of the hardest things. Babies lose heat during transfer from labour ward, theatre, or lower facilities. Even within the unit, if the mother removes the baby from KMC or delays changing wet clothes, the baby becomes cold*.*”* (KII6, nurse)

### Infection Prevention

Crowding, limited space, and decontamination-supply gaps were seen as drivers of sepsis.

> *“When the unit is crowded, it becomes difficult to separate infected and non-infected babies properly. We also need a consistent supply of gloves, sanitizers, feeding tubes, syringes, and antiseptics*.*”* (KII4, nurse)

### Drug and Equipment Availability

Oxygen, incubators, warmers, and CPAP were available but insufficient at peak demand, with occasional drug stock-outs delaying management.

> *“We have oxygen, some incubators, warmers, and CPAP, but when admissions are many, the equipment is not always enough for the number of babies… essential drugs are generally available, but there are moments when some items are delayed, or caregivers are asked to buy supplies*.*”* (KII1, pediatrician)

*“Sometimes we do not have enough incubator space or CPAP capacity. Laboratory services are available, but results may be delayed, so we often start treatment based on clinical judgment*.*”* (KII8, clinical officer)

### Referral Coordination

Weak pre-referral communication, incomplete notes, and poor transport produced delayed, cold, and unstable admissions.

> *“A small baby should not be transported cold. There should be clear referral protocols, including when to call, what to document, and how to maintain warmth during transfer*.*”* (KII9, unit manager)

### Monitoring, Timeliness, and Documentation

High workload slowed triage-to-admission care and constrained monitoring and record-keeping.

> *“Low-birth-weight babies need frequent monitoring, but when one nurse has many babies, it becomes difficult to check everything on time*.*”* (KII3, nurse)

*“You may be stabilizing one baby on oxygen, then another baby arrives from theatre or maternity. In such situations, prioritization is done, but some care processes may be delayed*.*”* (KII5, nurse)

### Caregiver and Transport Barriers

Limited recognition of danger signs, cultural beliefs, family decision-making, distance, and transport cost delayed care-seeking.

> *“Some families wait for permission from the father or elder. Transport cost is also a factor*.*”* (KII7, clinical officer)

### Integration: How Care Conditions Translate Into Outcomes

The quantitative and qualitative strands converge. The two care conditions statistically associated with severe outcomes—shortage of essential drugs/feeds and referral/outborn status—are precisely the conditions providers described as most disruptive: inconsistent commodity supply that delays management, and a poorly coordinated referral system that delivers cold, unstable, or undocumented neonates. The counter-intuitive warm-chain association is explained by the qualitative account that thermal support is concentrated on the smallest, sickest infants, and by the structural threats providers named (incubator shortages, congestion, and heat loss during transfer) that make thermal protection fragile precisely when it matters most. Near-universal skilled-personnel presence at admission, with no association to outcome, is consistent with providers’ distinction between the presence of staff and the adequacy of specialised training, equipment, and workload capacity. Table 5 maps each care condition to its integrated interpretation and the corresponding nursing-practice lever.

**Table 5:**
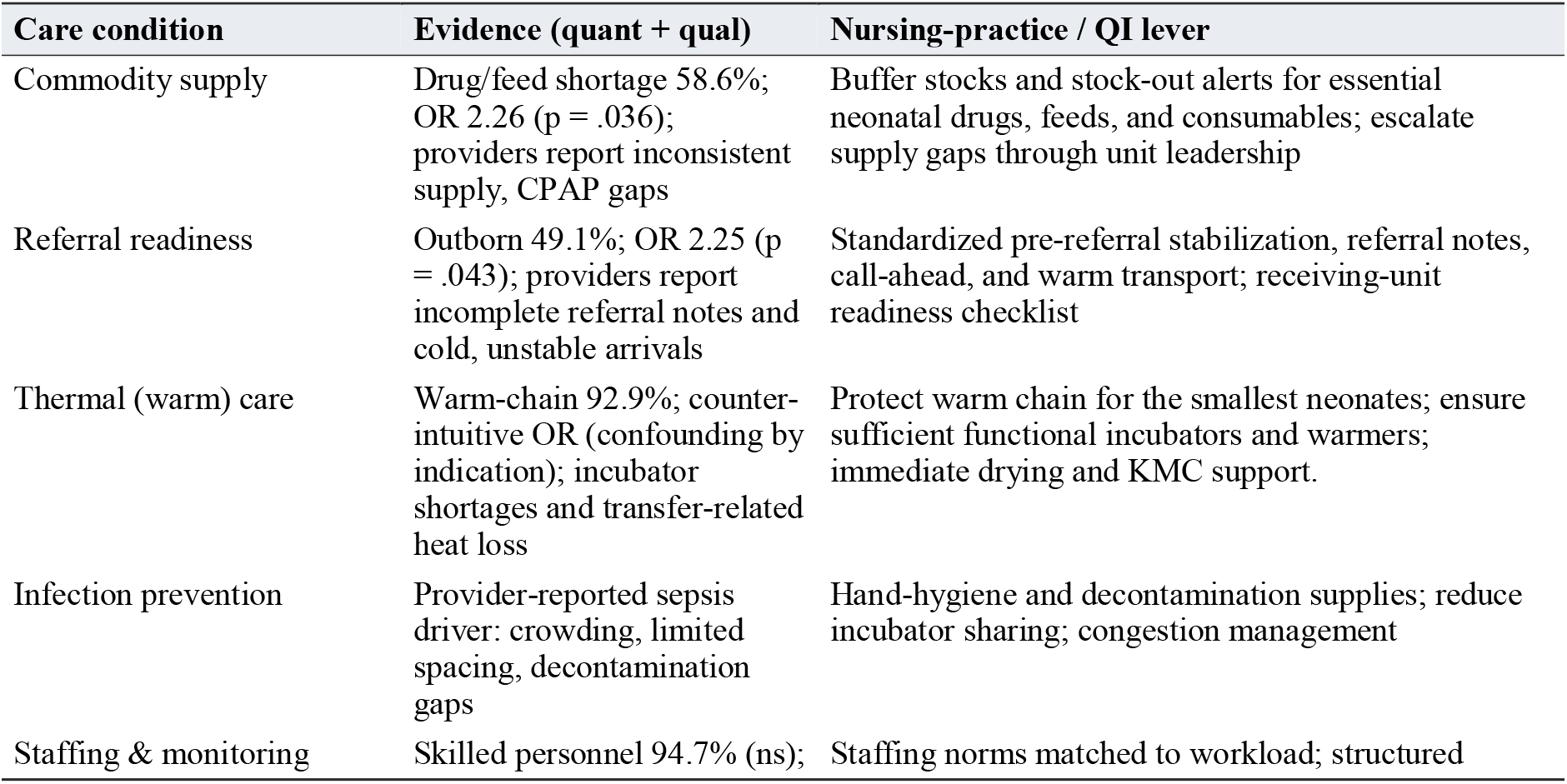
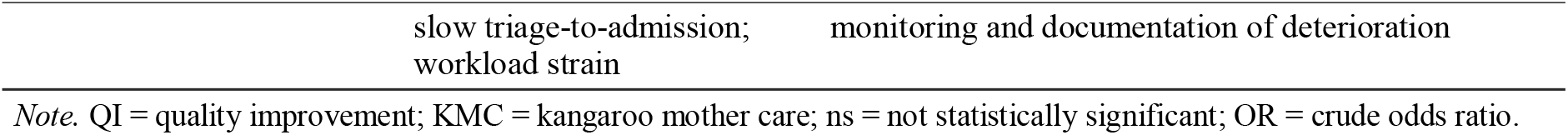
Integrated Nursing-Practice Implications by Care Condition.

Across all nine interviews, an overarching pattern connected the seven themes: adverse outcomes were seldom traced to a single failure, but to the interaction between bedside clinical care and system readiness. Providers described staff as skilled and responsive, yet repeatedly constrained by workload, equipment shortfalls, referral gaps, and intermittent supplies. This cross-cutting account informed the practice model in Figure 1, which was conceptualized as three linked layers: system-readiness conditions and structural barriers—staffing, commodities, equipment, and referral coordination—act through bedside care processes—timeliness, warm-chain maintenance, infection prevention, monitoring, and documentation—to shape the severe adverse outcome, with quality-improvement levers positioned to strengthen both layers simultaneously. *“Improving LBW outcomes requires both clinical care and system support. Staff can do their best, but without enough personnel, equipment, and referral coordination, some preventable complications will continue*.*”* (KII9, unit manager)

**Figure 1.**
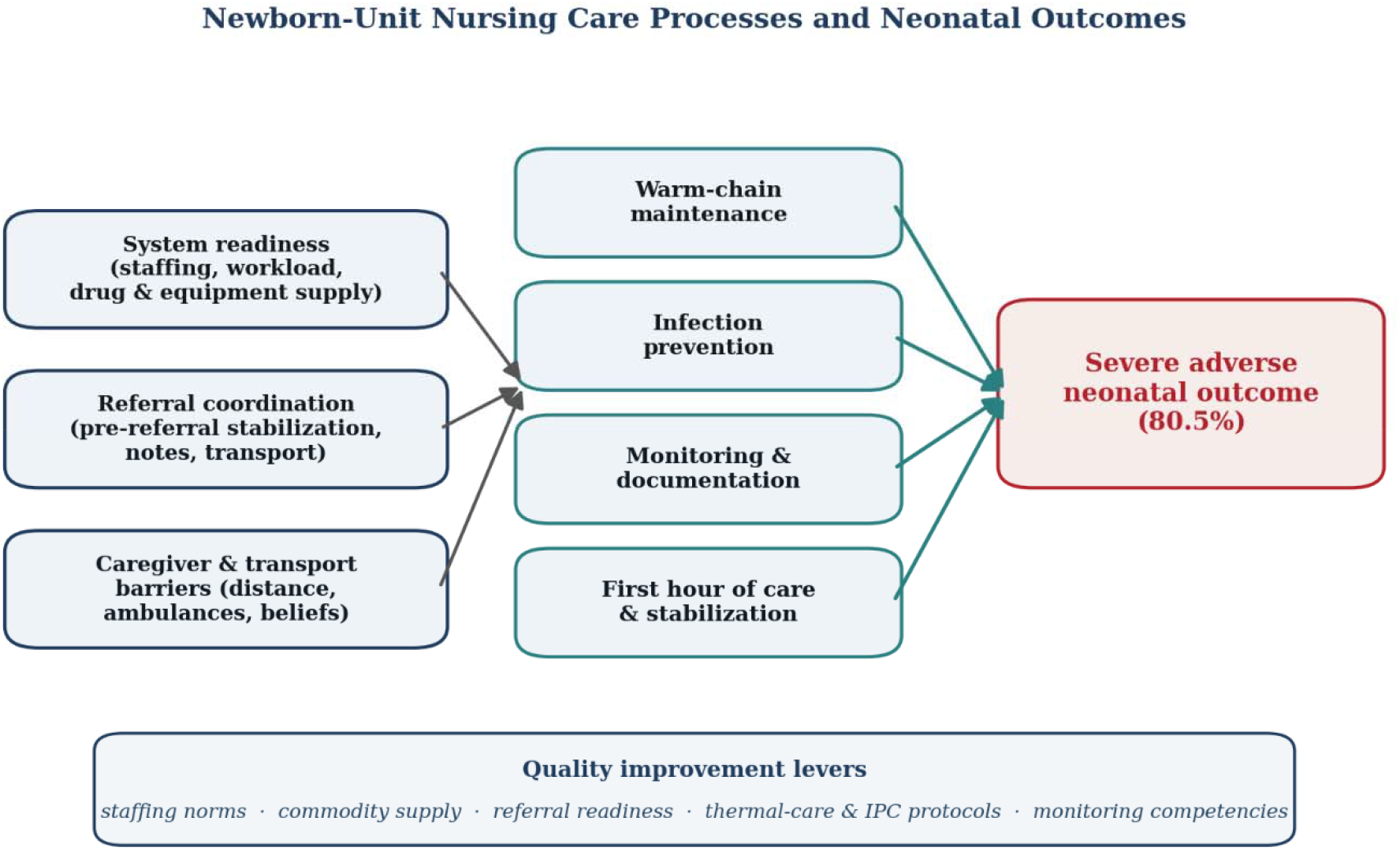
Practice model: newborn-unit nursing care processes and neonatal outcomes. *Note*. System-readiness conditions and structural barriers act through bedside care processes to shape the severe adverse outcome; quality-improvement levers target both layers.

## Discussion

This mixed-methods analysis shows that care-related conditions in the newborn unit are meaningfully linked to severe adverse outcomes among low-birth-weight neonates, and that provider perspectives explain the mechanisms behind those links. Two modifiable conditions— shortage of essential drugs/feeds and referral/outborn status—were each associated with roughly a doubling of the odds of a severe outcome, and both were the conditions providers described as most disruptive to timely, safe care. These findings reframe outcomes for vulnerable neonates as partly a function of system readiness and nursing-care conditions, not biology alone (WHO, 2022).

The association between referral/outborn status and severe outcomes is consistent with a large body of evidence that outborn neonates arrive colder, later, and sicker than inborn neonates, reflecting accumulated vulnerability across the referral pathway rather than transport distance alone (Kumar et al., 2020; Wainaina et al., 2023). Providers’ accounts of an unclear referral system, few ambulances, and poor pre-referral stabilization locate the actionable break-point at the interface between referring facilities and the receiving unit—an interface that nursing-led readiness checklists and standardized stabilization can strengthen.

The link between commodity shortages and worse outcomes echoes findings that stock-outs of essential newborn medicines and supplies undermine guideline-concordant care in Kenyan and regional newborn units (Murphy et al., 2018; Ministry of Health, Kenya, 2016). From a nursing standpoint, the practical lever is not only advocacy for supply but bedside systems—buffer stocks, stock-out alerts, and escalation—that reduce the clinical impact of intermittent supply.

The counter-intuitive warm-chain association warrants careful interpretation. Rather than implying that thermal care is harmful, it most likely reflects confounding by indication: the smallest, most premature neonates—those at highest baseline risk—are precisely those given the most intensive thermal support (Lunze et al., 2013). Provider narratives reinforce that the warm chain is under constant structural threat from power outages, incubator shortages, and congestion, meaning that near-universal documented warm-chain practice can coexist with real thermal instability. This mirrors cautions in the broader determinants literature that observational care-process associations can be distorted by severity (WHO, 2022).

That skilled-personnel presence at admission was near-universal yet unassociated with outcome underscores a distinction providers drew repeatedly: presence is not the same as capacity. Providers described gaps in NICU-specific training and in the ability to operate life-saving equipment such as CPAP and ventilators. This points education efforts toward competency-based training in neonatal resuscitation, thermal care, infection prevention, feeding support, and early recognition of deterioration, rather than headcount alone.

Infection prevention emerged as a central mechanism in the qualitative strand, with providers linking sepsis to crowding, shared incubators, and shortages of decontamination supplies. This is consistent with evidence that hospital-acquired infection is a leading and largely preventable contributor to neonatal mortality in low-resource units, where colonisation pressure is high and hand-hygiene compliance is difficult to sustain under heavy workload (Zaidi et al., 2005; Seale et al., 2014). For nursing practice, the actionable levers are concrete: reliable access to hand-hygiene and decontamination consumables, reduction of incubator sharing, and congestion management—each of which is a system condition rather than a fixed patient factor. Framing infection prevention as a care-process condition, rather than an individual behaviour, aligns responsibility with the unit-level resources and staffing that make safe practice possible.

The provider emphasis on slow triage-to-admission care and constrained monitoring points to the first hour after admission—the neonatal “golden hour”—as a high-yield window for LBW neonates. Structured stabilization and close monitoring in this period are associated with reductions in hypothermia, hypoglycaemia, and downstream morbidity (Sharma, 2017). Where staffing is thin, monitoring and documentation are the first tasks to be sacrificed during emergencies, so that early deterioration may go unrecognised. Strengthening structured observation, early-warning recognition, and documentation is thus not administrative overhead but a direct patient-safety intervention, and one squarely within nursing scope.

### Nursing education and quality-improvement implications

Taken together, the findings define a focused agenda for nursing education and quality improvement. Education should prioritise competency-based training—verified hands-on skill rather than attendance—in neonatal resuscitation, thermal care, infection prevention, feeding support, and recognition of early deterioration, including operation of CPAP and other life-saving equipment that providers reported being unable to use reliably. Quality-improvement work should target the conditions this study links to risk: buffer stocks and stock-out alerts for essential commodities, a receiving-unit referral-readiness checklist with pre-referral stabilization and warm transport, back-up power to protect incubation, and workload-matched staffing with protected time for monitoring and documentation. Because several of these levers sit at the facility and county level, they require unit leadership and health-system engagement alongside bedside change (English et al., 2017).

### Strengths and limitations

Strengths include the integration of measurable care-process indicators with provider narratives from multiple cadres, and the focus on modifiable conditions directly relevant to nursing practice. Limitations include the single-centre, cross-sectional design, which precludes causal inference; the small non-severe group (n = 33), which widens confidence intervals; the use of unadjusted associations, so that residual confounding—including confounding by indication—cannot be excluded; and reliance on documented care-process indicators, which may not capture the quality or timeliness of care. The qualitative strand, while spanning cadres, drew its verbatim illustrations chiefly from newborn-unit nursing experience. Findings should therefore be read as hypothesis-generating signals for quality improvement rather than definitive effect estimates.

## Conclusion

Care-related conditions—commodity supply, referral readiness, thermal care, infection prevention, staffing, and monitoring—are clinically modifiable levers that shape whether vulnerable low-birth-weight neonates stabilize or deteriorate. Even where individual care factors do not retain independent statistical significance after adjustment, they remain the conditions most amenable to change through nursing practice and system readiness. Strengthening bedside care processes and the systems that support them is therefore a direct and practical route to improving neonatal outcomes in county referral settings.

## Recommendations

### Practice

Standardize warm-chain maintenance for the smallest neonates (immediate drying, kangaroo mother care, and reliable incubator/warmer use with back-up power); reinforce hand-hygiene and decontamination routines and reduce incubator sharing; and implement structured monitoring for early recognition of deterioration in the first hour and beyond.

### Quality improvement

Establish buffer stocks and stock-out alerts for essential neonatal drugs, feeds, and consumables; adopt a receiving-unit referral-readiness checklist with pre-referral stabilization and warm transport; and set workload, protocol-adherence, and documentation targets for the newborn unit.

### Education

Prioritise ongoing, competency-based training in neonatal resuscitation, thermal care, infection prevention, feeding support, and recognition of early deterioration, including hands-on competence with CPAP and other life-saving equipment.

### Policy

At facility and county level, align nurse staffing norms with newborn-unit workload, secure consistent commodity supply, and invest in referral-system coordination and reliable power to protect thermal care.

## Data Availability

All data produced in the present study are available upon reasonable request to the authors

## Declarations

### Ethics approval and consent

Approved by the Kabarak University Research Ethics Committee (KUREC-170226; Ref. KABU01/KUREC/001/17/02/26) and licensed by NACOSTI (NACOSTI/P/26/4186446; Ref. 625542). Informed consent was obtained from all participants.

### Consent for publication

Not applicable; no individually identifiable data are presented.

### Availability of data

The de-identified datasets analysed are available from the corresponding author on reasonable request.

### Competing interests

The authors declare no competing interests.

### Funding

This research received no specific grant from any funding agency.

### Authors’ contributions

JC conceived and designed the study, collected and analysed the data, and drafted the manuscript. MK and MSS contributed to analysis, interpretation, and critical revision. EN contributed to design and revision. All authors read and approved the final manuscript.

## Acknowledgements

The authors thank the newborn-unit staff and administration of Kericho County Referral Hospital and the participating mothers and key informants.

## Notes

### Competing Interest Statement

The authors have declared no competing interest.

### Author Declarations

Kabarak University Research and Ethics Committee (KUREC-170226; ref. KABU01/KUREC/001/17/02/26) National Commission of Science, Technology and Innovation (NACOSTI/P/26/4186446)

